# *Boosting Everyday Movement*: Co-designing a digital micropatterns intervention with socioeconomically diverse UK and Australian Women

**DOI:** 10.1101/2025.10.29.25339035

**Authors:** Maria Bissett, Nicholas A. Koemel, Matthew Ahmadi, Kristina Atsiaris, Sam Liu, Cecilie Thøgersen-Ntoumani, Cindy M Gray, Jason Gill, Emmanuel Stamatakis, Gemma Ryde

## Abstract

**Background:** Vigorous (VILPA) and moderate-to-vigorous (MV-ILPA) intermittent lifestyle physical activity refer to brief bouts of physical activity (<1 and <3 minutes, respectively) embedded in daily routines. Evidence suggests that 4–6 daily bursts of VILPA/MV-ILPA can significantly reduce the risk of cardiovascular disease and some cancers. These “micropatterns” of activity may offer a time-efficient and accessible alternative to structured exercise, particularly for women from socioeconomically diverse backgrounds who face intersecting barriers to traditional forms of physical activity. This study aimed to explore women’s perspectives and experiences of VILPA/MV-ILPA and co-design a micropatterns intervention to promote these behaviours among socioeconomically diverse women.

**Methods:** The study involved a series of three co-design workshops with women in Glasgow (N=19) and Sydney (N=31). Workshops incorporated participatory activities, education, training, discussion, and reflection to explore the concept of micropatterns, related facilitators and barriers and co-design the intervention. This study was guided by the Behaviour Change Wheel and MRC and 6SQuID intervention development frameworks. Data were audio-recorded, transcribed, and analysed using thematic framework analysis.

**Results:** Participants identified a range of barriers (e.g. concerns about ability and safety, low mood, sociocultural norms) and facilitators (e.g. adaptability, dual-purpose activities, minimal time commitment) to engaging in micropatterns. Following reflection on the barriers and facilitators, six modifiable factors were identified to be addressed in the intervention, these included: lack of knowledge and awareness, concerns about ability and safety, low mood and poor mental health, sociocultural norms and environmental constraints. Participants identified thirteen intervention components that utilized seven intervention functions (education, persuasion, training, environmental restructuring, modelling, incentivisation, and enablement) to promote VILPA/MV-ILPA activities. Participants emphasised the importance of educational content, social support, and inclusive delivery formats (e.g. short videos, visual materials). Terminology such as “Mindful Movement” and “Boosting Everyday Movement” were preferred over technical acronyms and jargon.

**Conclusions:** The final intervention involved a six-week programme of education, training, goal setting and VILPA/MV-ILPA tracking. Due to the popularity of social components and mixed perceptions of the accessibility of digital technology, the intervention was designed with three different delivery mechanisms:1) a smartphone application, 2) a smartphone application and a wearable device (e.g. Fitbit) and 3) workshops, a smartphone application and a wearable device. With further testing, this co-designed intervention could offer a feasible approach to promoting physical activity micropatterns among women from diverse socioeconomic backgrounds.

## Introduction

Traditional exercise guidelines often recommend engaging in at least 150 minutes of moderate-to-vigorous physical activity (MVPA) per week (1). However, a substantial proportion of adults fail to achieve these targets, often due to time limitations and competing lifestyle demands (2, 3). Vigorous Intermittent Lifestyle Physical Activity (VILPA) is characterized by brief bouts of high-intensity movement integrated into existing daily routines (4–6). Unlike structured exercise programs, VILPA emphasises the intensification of everyday activities such as stair climbing, carrying heavy shopping, or brisk walking for transport performed in short bursts lasting <1 min (4–6). By embedding activity into routine tasks, VILPA has the potential to address key barriers to regular physical activity such as finding dedicated time for structured exercise.

Emerging wearable based evidence (4–8) has shown that these short bouts of vigorous activity is associated with improvements in cardiovascular health and mortality risk (4–8). The concept of VILPA is grounded in studies on high-intensity intermittent exercise, which demonstrate that even short durations of vigorous movement can yield substantial cardiovascular benefits, particularly among women (4–8). A large-scale cohort study involving 25,241 non-exercising UK adults demonstrated that accumulating one- to two-minute bouts of VILPA performed approximately three times per day, was associated with a 40% reduction in cancer-related mortality and an almost 50% reduction in mortality from cardiovascular disease (4). Recent evidence has highlighted that VILPA may be particularly beneficial for women, where 3.4 minutes of VILPA per day was associated with a 45% lower risk of overall major adverse cardiovascular events and a 67% lower risk of heart failure (5). This research highlights that even individuals who do not engage in traditional exercise can benefit from incorporating VILPA activities (e.g. outdoor work, heavy household chores, very fast walking or a burst of running (9)) into their daily routines. Studies have also reported analogous cardioprotective findings for moderate to vigorous intermittent lifestyle physical activity (MV-ILPA), undertaken in bouts up to 3 minutes in duration (9). MV-ILPA activities may include walking briskly, household chores and walking carrying light objects (9). These brief intermittent behaviours are collectively known as ‘*micropatterns*’ of physical activity.

As only 20% of adults regularly participate in leisure-time exercise, with incidental movement being the predominant source of physical activity for the majority of adults (10), micropatterns could offer a promising and feasible way to improve health among populations who face intersecting barriers to structured forms of leisure-time activity (e.g. gym, sports and exercise classes) including women and those from lower socioeconomic backgrounds (11). While prior research has begun to explore barriers and facilitators to VILPA/MV-ILPA (12–14) and tested small-scale interventions to promote it (15) studies have typically focused on middle-aged and older adults in Australia. Context is key in the development of interventions (16, 17), therefore to develop strategies to promote micropatterns in different populations and sub-groups (e.g. women), further research is needed to explore the unique perspectives and experiences of target groups. Thus, this study aimed to explore context-specific perspectives and experiences of VILPA/MV-ILPA and co-design a micropatterns intervention with women from socioeconomically diverse backgrounds in Glasgow and Sydney.

## Methods

### Study design

This study is a collaboration between the University of Glasgow and the University of Sydney and forms part of a phased approach to the co-design and feasibility testing of an intervention to promote micropatterns among socioeconomically diverse communities. This paper presents the findings from Phase I: a series of three co-design workshops with community members in Glasgow, Scotland and Sydney, Australia. The workshops involved a range of interactive activities, education, discussion and reflection to explore the concepts and terminology of VILPA/MV-ILPA, and co-design an intervention guided by COM-B and the Behaviour Change Wheel (18, 19). While the discussions were kept open to explore a range of intervention types, recent evidence has suggested that smartphone applications could be a promising component of an intervention to promote micropatterns (20), therefore participants were also explicitly asked to reflect on digital intervention components.

The co-design workshops were held in person (Glasgow and Sydney) and as hybrid sessions (Sydney only) between March and August 2025. During this period, continual meetings were held between the two countries to agree on and refine protocols to ensure consistency between groups.

### Ethics Statement

This research was approved by the University of Glasgow, College of Medicine, Veterinary & Life Sciences Ethics Committee (200240234) and the University of Sydney, Human Research Ethics Committee (HEOO1729). Prior to participation, interested participants in each setting were sent a detailed information sheet (by email or post). They were then phoned by a member of the research team, who provided further information on the study and answered any questions participants may have. All participants provided written informed consent at the beginning of the first workshop they attended.

### Participants, Recruitment and Screening

This study aimed to recruit 20-30 women in each context who would participate in three co-design workshops. The sample size was determined by the inherent nature of qualitative research, whereby the sample should be large enough to generate rich data while small enough to allow for deep, thorough analysis (21). It was also determined through consideration data adequacy (21): for example, the sample size per workshop supports the exploration of individual experiences and the provision of tailored communication and support as well as generating common group perspectives on micropatterns and intervention design.

Due to known socioeconomic inequalities in physical activity research and intervention design (22), recruitment strategies were prioritised in areas of higher deprivation. Researchers in each setting spent a significant period of time building and strengthening connections with local community organisations and visiting and posting fliers in community venues (e.g. local libraries, places of worship, housing associations and foodbanks). Posters were also shared on social media including via paid targeted Facebook advertisements.

Eligible participants were women aged 30-75 years, who self-reported little-to-no physical activity and were willing to and able to use smartphone apps or wearable technology. To screen for participants physical activity levels, interested participants were asked a series of questions adapted from the Global Physical Activity Questionnaire (GPAQ, (23)) which details domain-specific amounts of physical activity including leisure time, occupation, and transport. Prior to consent, potential participants were called and underwent screening to confirm eligibility. For health and safety purposes, participants were also screened using the Physical Activity Readiness Questionnaire (PARQ+) (24) where participants who answered yes to any of the questions were advised to seek approval from a health professional before taking part in the study.

### Frameworks for Intervention Development

The co-design of the intervention was informed by the Behaviour Change Wheel (BCW) developed by Michie and colleagues as a systematic way to design behaviour change interventions (18, 19). The BCW is formed from an inner circle composed of the COM-B model, a middle circle detailing Intervention Functions and an outer circle of Policy Categories. COM-B is a commonly used model that theorises that an individual must have the one or a combination of factors that provide them with the capability (C), opportunity (O) and motivation (M) to change a behaviour. This includes both the physical (e.g. physical ability) and psychological forms (e.g. knowledge and understanding) of capability, social and physical opportunities and automatic and reflective forms of motivation. By providing the intervention functions outside of the COM-B model, the BCW helps researchers and practitioners map behavioural determinants to targeted strategies, thus ensuring interventions are both theoretically grounded and practically applicable (18, 19).

The study was also guided by the six steps in quality intervention development (6SQuID) model (25) and the development phase of the MRC framework for developing and evaluating complex interventions (16). Specifically, the co-design workshops contributed to Steps 1-4 of the 6SQuID guide: 1) defining and understanding the problem and it’s causes; 2) identifying which causal or contextual factors are modifiable; 3) deciding on the mechanisms of change and 4) clarifying how they will be delivered (25). Intervention co-design workshops were also designed to address key action points from the MRC framework (16, 17). For example, this study drew on existing evidence from previous VILPA studies, involved community members in Glasgow and Sydney to understand contextual barriers and facilitators. It also centred participants as key stakeholders in the design of the intervention to inform the development of an intervention programme theory (16, 17).

### Data Collection

#### Demographic Data

At Workshop 1, participants were asked to complete a short demographic and health-related survey including age, ethnicity, socioeconomic status, postcode, household income range, number of dependents, and highest education level.

Postcodes were then used to determine area-based deprivation. In Scotland, this was done using the Scottish Index for Multiple Deprivation (SIMD) scores which rank areas across Scotland from 1 to 10 according to different indicators of deprivation, 1 being the most deprived and 10 being the least deprived (26). In Sydney this was done using Socio-Economic Indexes for Areas (SEIFA) which similarly provides area-based rankings in Australia based on different socioeconomic indexes-Level 1 captures the least advantaged areas and Level 5 captures the most advantaged areas (27).

Participants were also asked to rate their overall health using the EQ5D VAS scale which asks participants to rate their general health on a scale from 0 ’worst imaginable health state’ to 100 ‘best imaginable health state’ (28, 29).

### Co-Design Workshops

The workshops were designed to be as accessible as possible. Thus, although the same workshop protocol was followed in Glasgow and Sydney, the delivery of each session was tailored to each context. In Glasgow, two sets of workshops were delivered in person only, one at community centre in a deprived area in the South of Glasgow (Workshop A) and another held on campus at the University of Glasgow in the West End (Workshop B). The workshops were scheduled on different days (Tuesday and Wednesday) to offer participants different sessions that they could attend or swap between around their schedules or commitments. In Sydney, one set of hybrid workshops were held at the University of Sydney, Westmead Innovation Centre and online for those not able to attend sessions in person. Participants were reimbursed for their travel expenses, received refreshments and were provided with £60 ($120) of shopping vouchers for their involvement.

The objectives and key activities of each workshop are summarised in Table 1. Each workshop had one lead facilitator (MB in Glasgow and NK in Sydney) who were supported by co-facilitators each week. Workshops 1-3 consisted of a range of

**Table 1:** Summary of co-design workshops 1-3 including objectives and key activities

| **Summary of Co-Design Workshop Objectives and Activities** |  |
| --- | --- |
| **Workshop 1: An Introduction to VILPA/MV-ILPA and Terminology** |  |
| **Summary Objectives** | **Key Activities** |
| • Introduce the concept of physical activity with focus on micropatterns, describing different intensities and the benefits.<br>• Develop an understanding of participant's perspectives on, and experiences of, VILPA/MV-ILPA with a focus on barriers and facilitators and terminology. | • Education and Discussion:<br>○ Overview of physical activity and terminologies,<br>○ What are VILPA/MV-ILPA and what are the benefits?<br>• Interactive activity using post it notes: What are the facilitators and barriers to VILPA/MV-ILPA?<br>• Small Group Work: Brainstorm new terminology for VILPA/MV-ILPA |
| **Workshop 2: Experience VILPA/MV-ILPA** |  |
| **Summary Objectives** | **Key Activities** |
| • Work with participants to identify opportunities for micropatterns in their daily life<br>• Work with participants to self-identify when they have reached a certain intensity/exertion when undertaking VILPA/MV-ILPA | • Co-create a list of opportunities for micropatterns<br>• Try a series of VILPA/MV-ILPA activities and measure intensity/exertion using talk test, heart rate monitors and rate of perceived exertion<br>• Reflection on trying VILPA/MV-ILPA |
| **Workshop 3: Co-Designing a VILPA/MV-ILPA Intervention** |  |
| **Summary Objectives** | **Key Activities** |
| • Co-design a feasible and sustainable micropattern intervention, including but not limited to the frequency, timing, and duration of bouts.<br>• Develop an understanding of what activities/devices (e.g. of consumer-grade wearables, smart-phone-technology, and educational materials) may be practical and promote adherence to a VILPA/MV-ILPA intervention | • Reflection on Trying VILPA/MV-ILPA at Home<br>• Education: Introduction to COM-B and Behaviour Change Theory<br>• Small Group Work: Use COM-B analysis to identify modifiable factors and prioritise key intervention components<br>• Small Group Work and Discussion: explore wider delivery mechanisms and concepts such as frequency, timing, and duration of bouts, usage of technology and return to terminology. |

**Table 2:**
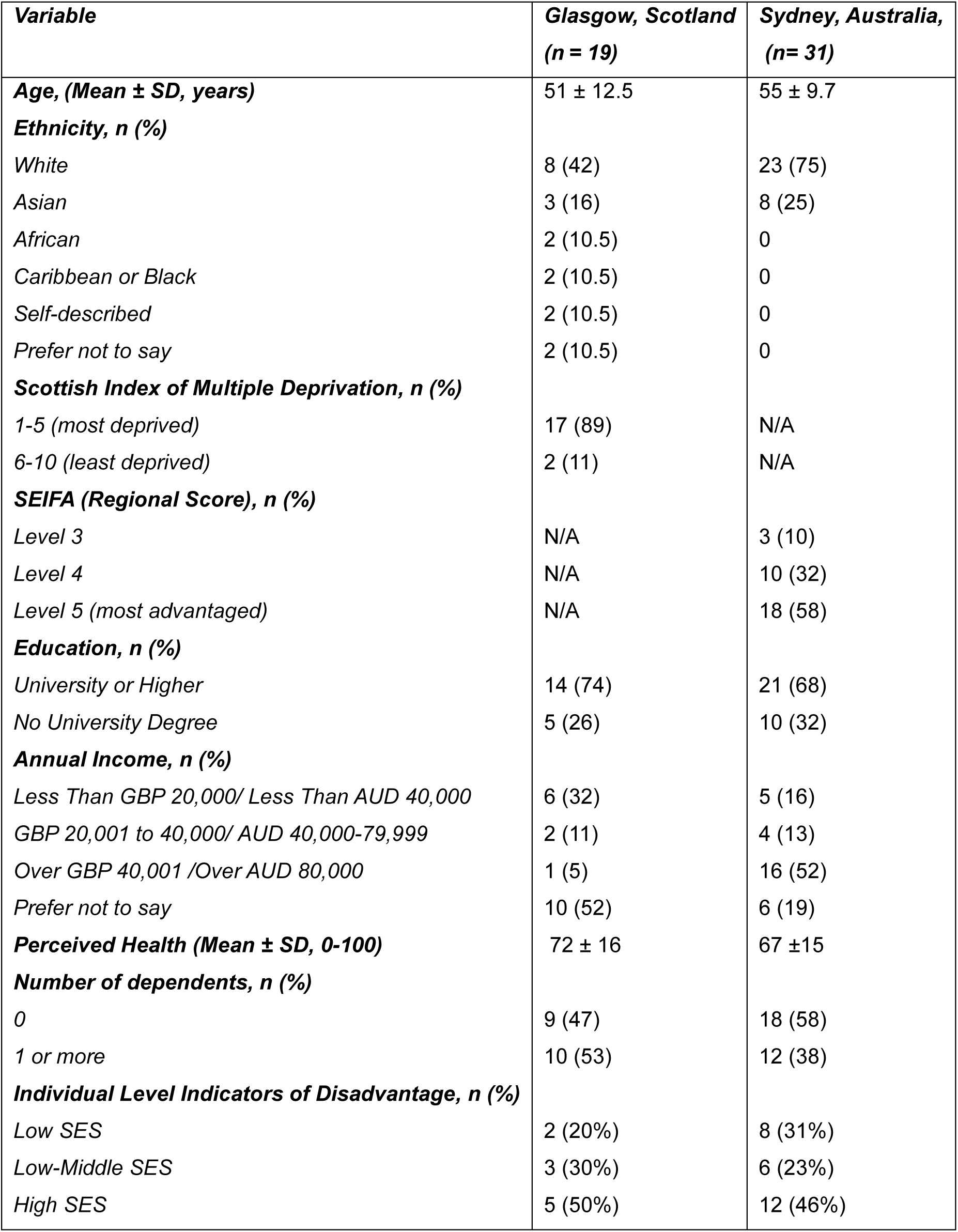
Summary of female demographics in Glasgow and Sydney including age, ethnicity, area-based deprivation score (SIMD Scotland and SEIFA Australia), level of education, annual income range, self-reported overall health score (0-100, 100 being best possible health), number of dependents and individual level indicators of socioeconomic disadvantage.

| **Variable** | **Glasgow, Scotland**<br>**(n = 19)** | **Sydney, Australia,**<br>**(n= 31)** |
| --- | --- | --- |
| **Age, (Mean ± SD, years)** | 51 ± 12.5 | 55 ± 9.7 |
| **Ethnicity, n (%)** |  |  |
| White | 8 (42) | 23 (75) |
| Asian | 3 (16) | 8 (25) |
| African | 2 (10.5) | 0 |
| Caribbean or Black | 2 (10.5) | 0 |
| Self-described | 2 (10.5) | 0 |
| Prefer not to say | 2 (10.5) | 0 |
| **Scottish Index of Multiple Deprivation, n (%)** |  |  |
| 1-5 (most deprived) | 17 (89) | N/A |
| 6-10 (least deprived) | 2 (11) | N/A |
| **SEIFA (Regional Score), n (%)** |  |  |
| Level 3 | N/A | 3 (10) |
| Level 4 | N/A | 10 (32) |
| Level 5 (most advantaged) | N/A | 18 (58) |
| **Education, n (%)** |  |  |
| University or Higher | 14 (74) | 21 (68) |
| No University Degree | 5 (26) | 10 (32) |
| **Annual Income, n (%)** |  |  |
| Less Than GBP 20,000/ Less Than AUD 40,000 | 6 (32) | 5 (16) |
| GBP 20,001 to 40,000/ AUD 40,000-79,999 | 2 (11) | 4 (13) |
| Over GBP 40,001 /Over AUD 80,000 | 1 (5) | 16 (52) |
| Prefer not to say | 10 (52) | 6 (19) |
| **Perceived Health (Mean ± SD, 0-100)** | 72 ± 16 | 67 ± 15 |
| **Number of dependents, n (%)** |  |  |
| 0 | 9 (47) | 18 (58) |
| 1 or more | 10 (53) | 12 (38) |
| **Individual Level Indicators of Disadvantage, n (%)** |  |  |
| Low SES | 2 (20%) | 8 (31%) |
| Low-Middle SES | 3 (30%) | 6 (23%) |
| High SES | 5 (50%) | 12 (46%) |

participatory activities, education, group discussion and reflection. Workshop 1, focused on an introduction to physical activity and the concept of micropatterns, facilitators and barriers and related terminology. In Workshop 2, participants were invited to volunteer to try VILPA and MV-ILPA activities to help understand the required intensity of vigorous and moderate-vigorous activity. In Glasgow, activities (e.g. speed walking, stair climbing) were done using nearby parks and greenspace where participants could make use of existing hills and outdoor staircases. In Sydney in person workshops, participants did stair climbing, brisk walking indoors and outdoors and replicated picking up household items using sports cones. These activities were encouraged for hybrid participants; however, they were also asked to adapt the activities to the environment they were in, including brisk walking outdoors or marching in place at tempo guidance. At in person workshops, volunteers could wear heart rate monitors to track the intensity of their movement, while other group members timed their activities, and helped to measure the intensity using talk tests and the rate of perceived exertion scale (30, 31).

Between workshops 2 and 3, the research team conducted an analysis of the transcripts from Workshops 1 and 2 using COM-B (18, 19). This analysis was then used to create a summary table of perceived facilitators and barriers that was shared with participants in Workshop 3. Participants were provided with a short presentation on COM-B and how it is used to inform behaviour change interventions. They were then asked to collaboratively sense-check the analysis so far and to identify the most important modifiable factors that should be focused on in an intervention to promote VILPA/MV-ILPA. This was followed by small group work to identify key intervention delivery mechanisms including a discussion on the components of a smartphone application.

### Data Processing and Analysis

Demographic and health data were processed and summarised descriptively in Excel. All workshops were audio-recorded, transcribed verbatim and pseudonymised by the research team (MB in Glasgow and KA in Sydney). Fieldnotes were written up using an observation proforma and online chats from the Sydney hybrid sessions were exported for analysis. All data were analysed using thematic framework approach (32). Following familiarisation, three Sydney transcripts (and accompanying online chats from the hybrid session) were independently coded by MB (Glasgow) and KA (Sydney) using Nvivo 14. The researchers then met online to review and discuss the coding and agree an initial framework for analysis. There were four broad themes that each contained a range of subthemes. The broad themes included: *Facilitators of VILPA/MV-ILPA, Barriers to VILPA/MV-ILPA, Designing the Intervention* and *Intervention Delivery Mechanisms*. MB then applied and refined the framework to code the remaining transcripts from Glasgow.

Framework matrices were constructed in Excel for each overarching theme and its subthemes. The subthemes were then summarised and mapped to the behaviour change wheel domains (18, 19). The analysis was reviewed and cross-checked throughout by researchers at Glasgow (MB and GR) and Sydney (NK and KA).

## Results

### Participants

Participant demographics, as shown in Table 1, were broadly similar in both settings. In Glasgow, participants were aged 51±12.5 years old on average, White European (42%), African, Caribbean or Black (21%) or Asian (16%), lived in deprived areas (89%) and were university educated (75%). Most participants had an average perceived health of 72 (100 meaning best possible health). In Sydney, participants were 55±9.7 years old on average, white (75%) or Asian (25%), university educated (68%) and had an average perceived health of 67. The majority of participants in Sydney lived in the most advantaged areas (58%) and many had with an income of over $80,000 AUD (52%). To understand individual-level socioeconomic status, a composite score was calculated using educational attainment and reported income with scores ranging from 1.0–2.5 classified as low, >2.5–3.5 as medium, and >3.5–5.0 as high. Using this scoring, there was diversity across socioeconomic status in Sydney (31% low, 23% medium and 46% high) and Glasgow (20% low, 30% medium and 50% high). However, it should be noted that 52% of participants in Glasgow preferred not to report their income.

In Glasgow, two people missed Workshop 1 but were provided with the slides and caught up at the beginning of Workshop 2. In Sydney, the majority of participants attended the workshops online (77% in Workshop 1, 76% in Workshop 2 and 78% in Workshop 3) and three participants missed Workshop 1.

### Intervention Co-Design

The results of the co-design workshops are presented according to Steps 1-4 of the 6SQuID guide for intervention development (25). Each of the three co-design workshops contributed to Step 1: understanding the causes of the problem i.e. not enough VILPA/MV-ILPA. Workshop 3 addressed the remaining steps: 2) identifying which causal or contextual factors are modifiable; 3) deciding on the mechanisms of change and 4) clarifying how they will be delivered.

#### Step 1. Understanding the causes: Barriers and Facilitators of VILPA/MV-ILPA

A recent scoping review suggested that an individual’s skill, environmental barriers and social influences were the key barriers to components of VILPA (13).

Furthermore, previous qualitative investigations with middle aged and older adults in Australia have identified barriers including negative perceptions of vigorous physical activity and the associated effort required, physical limitations and health constraints, environmental constraints such as poor weather, fear of injury and a lack of awareness of the concept (12, 14). Despite this growing body of work, due to the relative novelty of VILPA as a concept and the lack of evidence from a UK context and among women from lower socioeconomic backgrounds, it was fundamental to explore the context-specific barriers and facilitators with participants in Glasgow and Sydney in the present study.

### Barriers to VILPA/MV-ILPA

In the present study, there were seven key barriers that could prevent engagement in VILPA/MV-ILPA. These include *Ageing and Physical Decline, Lack of Knowledge or Awareness of VILPA/MV-ILPA, Concerns about Ability and Safety, Low Mood or Poor Mental Health, Perceived Energy and Effort, Sociocultural Barriers and Environmental Constraints*. In Table 3, the themes are mapped to the COM-B behavioural model and example quotes are provided from Glasgow and Sydney.

**Table 3.** A Summary of the perceived barriers to VILPA/MV-ILPA mapped to COM-B with examples and representative quotes from Glasgow and Sydney.

| Theme<br>(COM-B domain) | Examples | Representative Quote:<br>Glasgow | Representative Quote:<br>Sydney |
| --- | --- | --- | --- |
| **Ageing and Physical Decline**<br>(Physical Capability) | Loss of strength<br>e.g. due to menopause or arthritis<br>Fear of falling, especially on stairs or uneven pavements | *"Because I walked in the house mostly without a stick. I use it sometimes, but when I'm outside, I use my stroller, the condition of pavements. The fact that in your mind, I don't want a fall at my age, I don't want a fall..."*<br>Participant<br>6_Glasgow_Workshop 1A | *"Because my arthritis was really bad, it's like I just can't move in the way that I used to... [My] strength is not as good"*<br>Participant<br>27_Sydney_Workshop 1 |
| **Lack of Knowledge or Awareness of VILPA/MV-ILPA**<br>(Psychological Capability) | Concept of VILPA/MV-ILPA sits counter to traditional conceptualisations of exercise and PA | *"Like it never occurred to me like this is so short. Just walk it faster, you know? [...] But the other thing is it's like; how do you speed up?"*<br>Participant<br>15_Glasgow_Workshop 3B | *"After last week's session I shared what we talked about with 2 friends, and we all agreed that being informed has changed what we did this week. You know we've all been conditioned to think you have to go to the gym for an hour and work out and stress yourself and get your heart rate up, etc."*<br>Participant<br>29_Sydney_Workshop 2 |
| **Concerns about Ability and Safety**<br>(Automatic Motivation) | Worry about heart rate spikes during movement<br>Fear of injury from sudden bursts of activity | *"I tried to upgrade [MV-ILPA] to VILPA, but you know, I was afraid to do that because I don't want to do something all of a sudden and then I would have those cramps and all the like and like pushing yourself to extremes."* | *"So, I sometimes worry, you know, like, if I did some stretches, I might be stretching wrong"*<br>Participant<br>16_Sydney_Workshop 1 |
|  |  | Participant<br>4_Glasgow_Workshop 3B |  |
| **Low Mood and Poor Mental Health**<br>**(Automatic Motivation)** | Depression, low mood, and emotional fatigue<br>Dissonance between intention and action | *"I became quite grumpy in life in general. So, and it was actually quite shocking to realise that some of your emotions affect whether you can be bothered to do something or not"*<br>Participant<br>15_Glasgow_Workshop 3B | *"I might have a bad day and do nothing"*<br>Participant 6_Sydney Workshop 2 |
| **Perceived Energy and Effort required**<br>**(Automatic Motivation)** | Effort required to do VILPA<br>Tiredness from work or responsibilities<br>"Laziness" | *"But then, like, I feel so tired and I'm like if this is something I have to do every day...don't think I'll be running like this like every day. So, I did away with that, the VILPA. I now concentrated on that MV-ILPA part"*<br>Participant<br>13_Glasgow_Workshop 3B | *"Being worn out too quickly is off-putting"*<br>Participant<br>6_Sydney_Workshop 2 |
| **Sociocultural Barriers**<br>**(Social Opportunity)** | Embarrassment or wariness around exercising in public<br>Cultural norms discouraging women from running or cycling<br>Dress code concerns affecting comfort and confidence | *"Well for me, because you know, because of my dress code, because the way I am, I was a bit wary of what people would think that why she's just running in the street"* Participant<br>17_Glasgow_Workshop 3B | No explicit data about barriers but do discuss cultural adaptations e.g. language in Workshop 3 |
| **Environmental Constraints**<br>**(Physical Opportunity)** | Poor weather<br>Lack of infrastructure (e.g. stairs, safe walking paths)<br>Dirty or unsafe environments | *"Being unable to go out in winter myself, you know, it's because [dark evenings] can be scary, very hard to go out"*<br>Participant<br>2_Glasgow_Workshop 1A | *"When it's cold we don't want to exercise".*<br>Participant<br>29_Sydney_Workshop 1 |

One of the key barriers to VILPA/MV-ILPA was *ageing and physical decline* which was often linked to health conditions or fear of injuries due to lower strength and mobility. C*oncerns about ability and safety* were not only linked to age but also the intensity of the activity and environmental infrastructure. In Glasgow, several younger participants highlighted worries about the impact of sudden intense movement on their bodies. This was particularly noticeable once they had tried doing VILPA at home between Workshops 2 and 3. Another important barrier appeared to be *a lack of knowledge or awareness of VILPA/MV-ILPA*. Participants in Glasgow and Sydney revealed the concept of VILPA/MV-ILPA could sit counter to traditional conceptualisations of exercise which prioritises extended periods of structured movement. The need for more knowledge about VILPA/MV-ILPA also appeared to be most apparent to participants after they had received some education themselves during the workshops. The *perceived energy and effort* required to do activities like VILPA/MV-ILPA was also a barrier for many participants. Participant 6 in Sydney Workshop 2 highlighted that *"being worn out too quickly"* would be off-putting while participants in Glasgow Workshop 1A said they wouldn’t have done VILPA/MV-ILPA because they were too tired from work [Participant 6] or due to “*laziness”* [Participant 7]. Similarly, after trying VILPA/MV-ILPA in Glasgow, several participants suggested that VILPA was too intense, but MV-ILPA might be more achievable. This is an important factor to consider when deciding what the starting point of the intervention might be.

*Environmental constraints* were another barrier to VILPA/MV-ILPA for many participants in both Glasgow and Syndey. Several participants suggested that the weather could negatively impact their motivation. As noted by participant 29 in Sydney Workshop 1, *“when it’s cold we don’t want to exercise”.* Additionally, some participants highlighted that not having physical or social opportunities in their environment (e.g. sedentary job, lack of hills, stairs, or family members to play with) could make increasing the intensity of their movement more difficult. In Glasgow, sociocultural barriers were discussed more explicitly than in Sydney, where participants from minoritised ethnic backgrounds highlighted sociocultural norms could put them off certain activities (e.g. cycling) or make them more aware of their presence when running in the street in their traditional dress. Finally, participants highlighted that their motivation to do physical activity could be negatively impacted by their *low mood or poor mental health*.

### Facilitators of VILPA/MV-ILPA

Several facilitators were identified that could support engagement in VILPA/MV-ILPA. These included: *Adaptability and Choice; Dual-Purpose Activities; Community and Social Support, Enjoyment and Fun; Mental Health Benefits; Valuing Physical Health Benefits; Routine, Prompts, and Scheduling; Personal Determination; Valuing Short, Manageable Movement; Knowledge and Understanding, and a Conducive Environment.* These themes are mapped to the COM-B behavioural model and example quotes are provided from Glasgow and Sydney in Table 4.

**Table 4.**
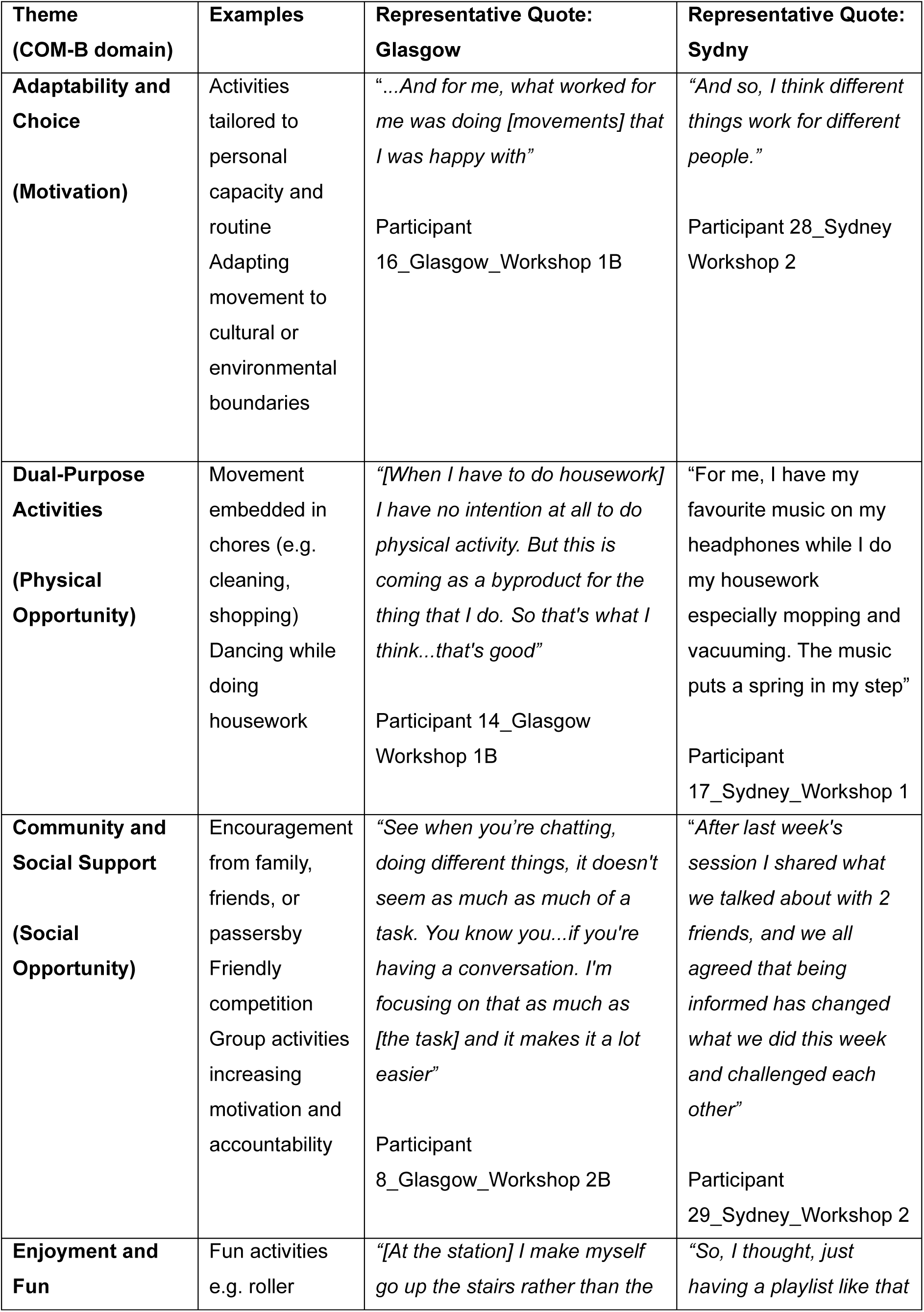
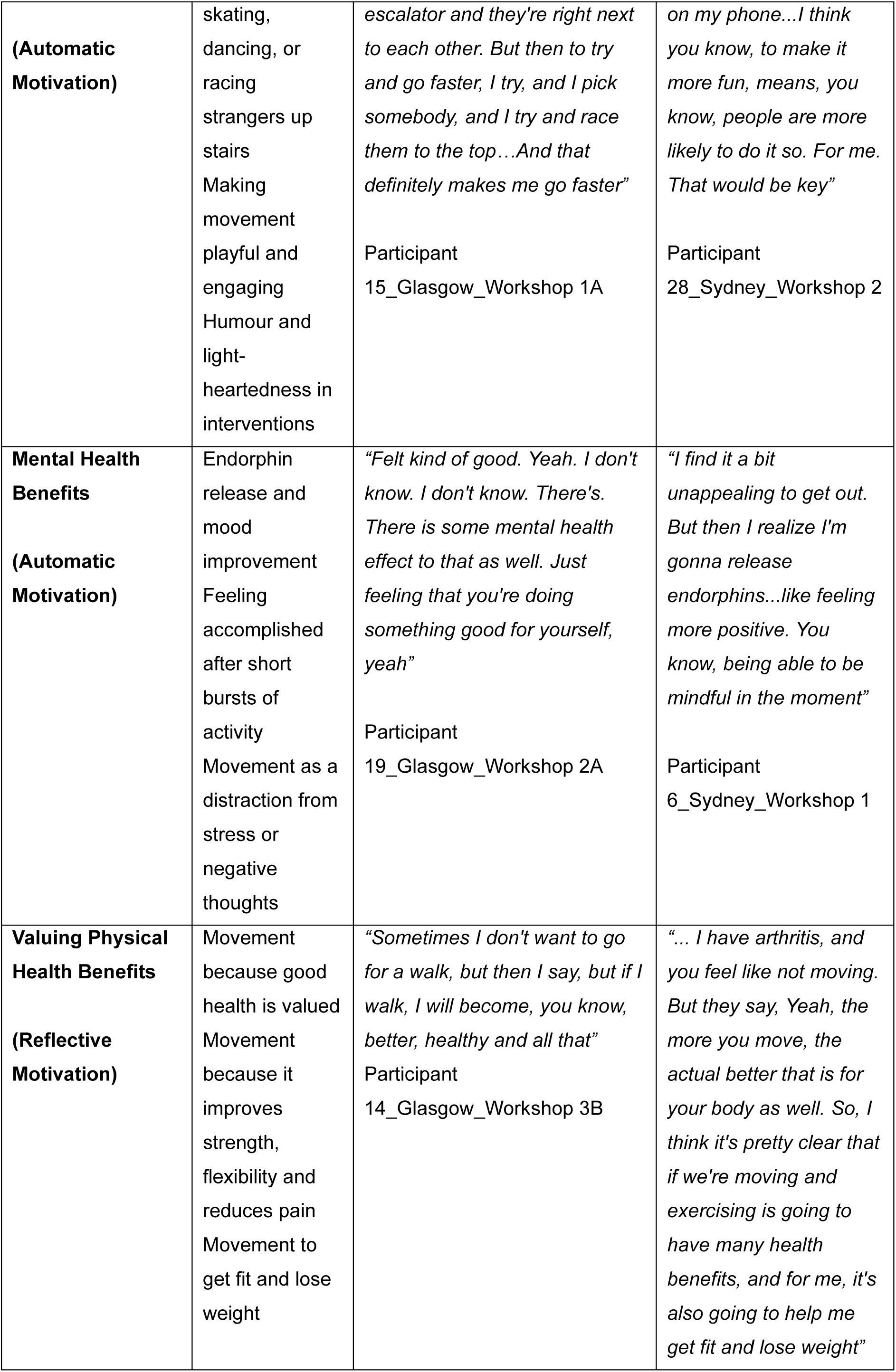
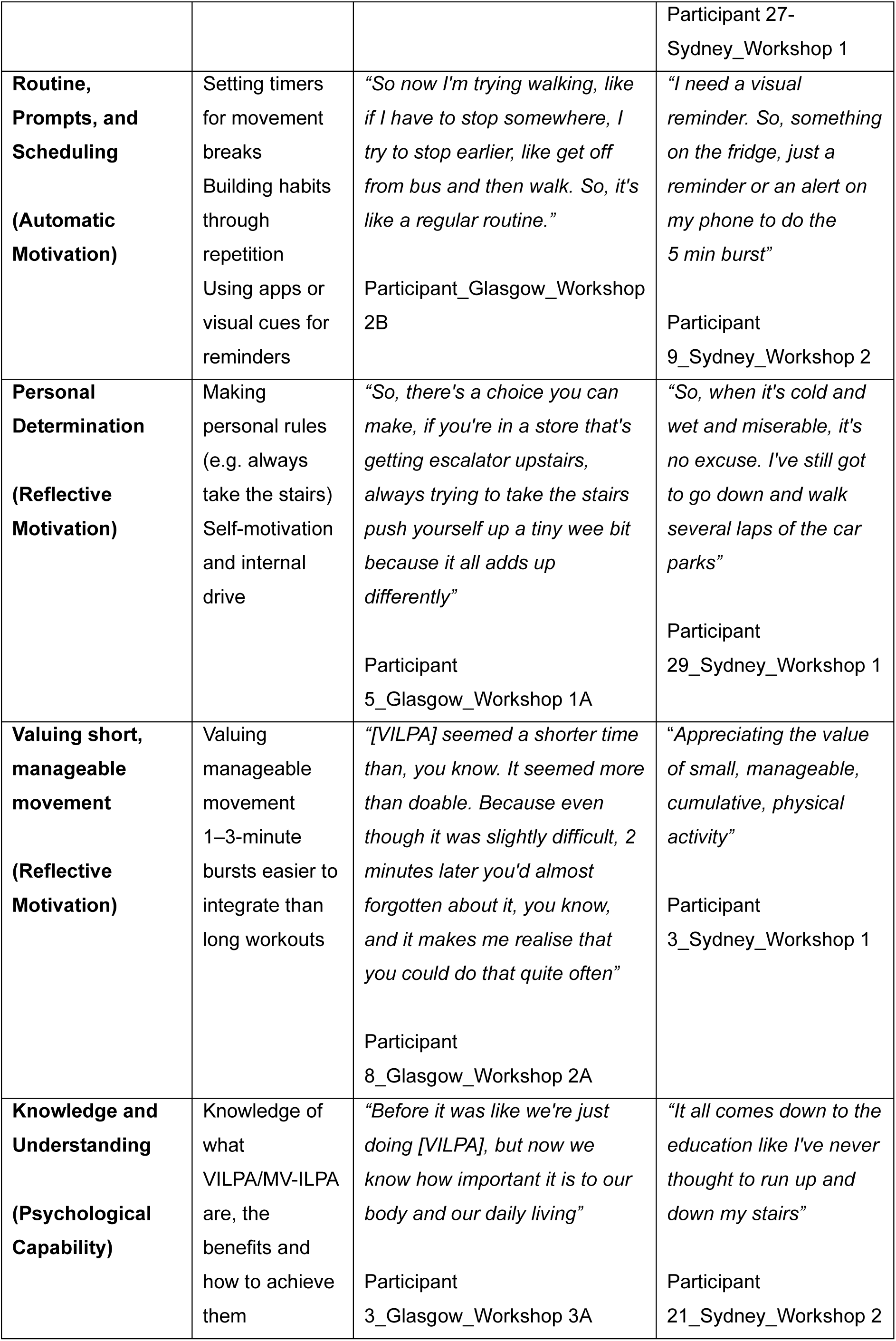
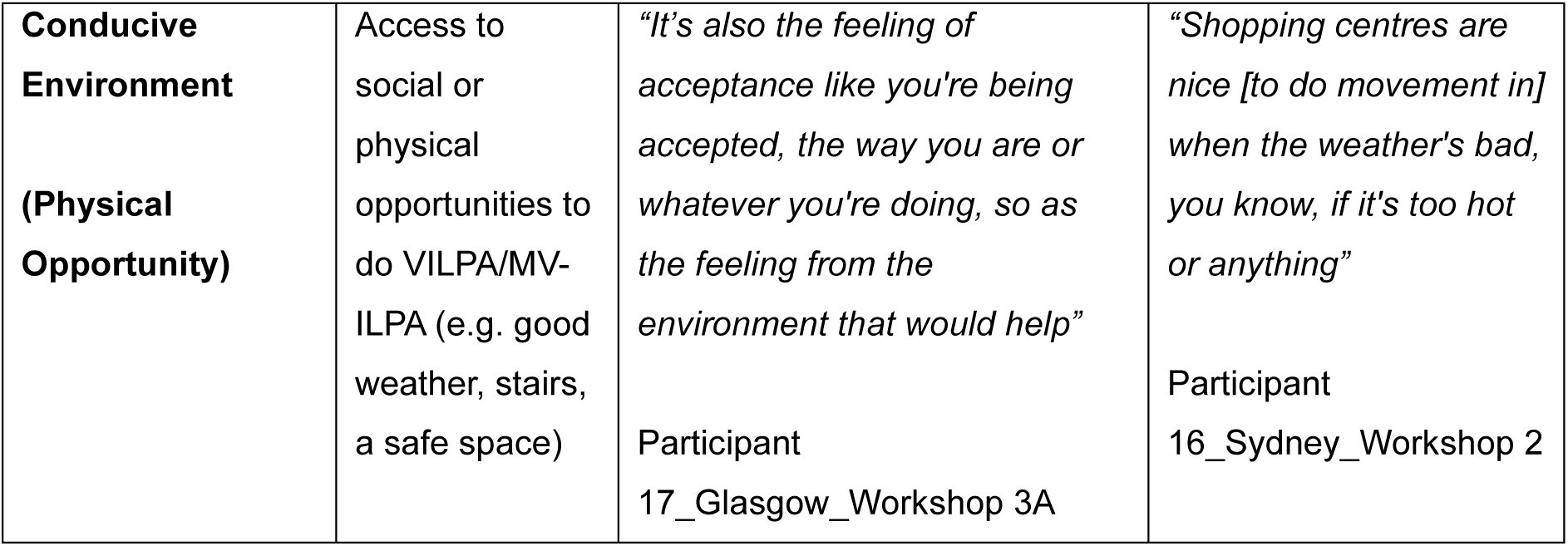
A summary of the perceived facilitators of VILPA/MV-ILPA mapped to COM-B with examples and representative quotes from Glasgow and Sydney.

A common theme was the importance of *adaptability and choice*, specifically participants in both contexts highlighted VILPA/MV-ILPA was appealing in that it could be tailored to different routines, preferences and cultural contexts. Another key facilitator was the *dual-purpose* nature of VILPA/MV-ILPA where movement is embedded in daily chores and participants praised the idea that they could achieve health and lifestyle benefits simultaneously. *Community and social support* also emerged as a strong motivator, with participants valuing encouragement and shared experiences with friends and family. *Enjoyment and fun* were similarly key, with playful approaches like creating imaginary games (e.g. racing strangers up the stairs), using music or dancing during housework enhanced their motivation to increase the intensity of daily movements. Participants in Glasgow and Sydney also highlighted that the *mental health benefits of VILPA/MV-ILPA*, such as mood improvement and stress relief could also encourage them to take part. Many participants also *valued the long-term health benefits* that they could achieve by doing VILPA/MV-ILPA as well as more immediate benefits like improved muscle tone.

Another important theme was *establishing routine and prompts*, where tools like visual reminders or app alerts, helped build habits, while the nature of VILPA/MV-ILPA as *short, manageable bursts of movement* made the concept feel achievable for many. Finally, *knowledge and understanding* of VILPA/MV-ILPA as a concept and a *conducive environment* that included safe spaces, infrastructure (e.g. stairs) and good weather were fundamental.

#### Step 2. Identifying what factors are modifiable

In Workshop 3, the research team and participants reflected on all the barriers and facilitators that had emerged in Workshops 2 and 3 and worked to identify the factors that were most important to modify with consideration of the study’s limited capacity and resources. In both contexts, the importance of knowledge and awareness across multiple levels and the adaptability and do-ability of VILPA/MV-ILPA were key. In Glasgow, participants also emphasised the importance of safety. These factors were then considered among the teams in Glasgow and Sydney with consideration of capacity and resources. *Knowledge and awareness, perceptions of energy and effort, concerns about ability and safety,* and *low mood and mental health* were agreed to be readily modifiable. Factors like the *physical and social environment*, could be partially modifiable with certain strategies (e.g. providing a list of different opportunities for and examples of VILPA/MV-ILPA). Finally, *ageing and physical limitations* were viewed as not modifiable within the scope of the intervention.

#### Step 3. Identify the change mechanism

Participants in Workshop 3 identified four broad areas that the intervention should focus on: *Education and Awareness; Social Support and Community Building; Goal setting; and Tracking and Gamification.* These themes are summarised briefly below and are mapped to Behaviour Change Wheel Intervention functions in Table 5. In Table 6, the intervention components that were included in the final intervention are summarised and mapped to the intervention functions and modifiable factors they address.

**Table 5:** Summary of key themes that emerged during the intervention co-design activities mapped to behaviour change wheel intervention functions. Representative quotes from Glasgow and Sydney are provided.

| Theme<br>(Intervention Function) | Examples | Representative Quote: Glasgow | Representative Quote: Sydney |
| --- | --- | --- | --- |
| **Education and Awareness**<br>(Education, Training and Persuasion) | Explaining what VILPA/MV-ILPA are, the health benefits, the accessibility and how to do it. | *“What do others need to know? We said: they need to know that these activities help reduce the risk of some diseases like cancer, and then they also need to know that there are a variety of everyday activities to choose from, so it's easy for them to do what they like. Then they should also know that they can be tailored to one's ability”*<br>Participant<br>19_Glasgow_Workshop 3B | *“So, the message. It's most important rather than introduce the [VILPA/MV-ILPA], if you like to convince people about how easy it is adopting their micropatterns into the life rather than try to convince benefits first”*<br>Participant<br>5_Sydney_Workshop 3 |
| **Social support and community building**<br>(Modelling and Environmental Restructuring: Social) | Buddy systems and walking groups. Online forums for sharing progress and encouragement<br>Accountability through shared goals | *P4: It's better if everybody gets involved, we have a giggle at the same time.*<br>*P7: Aye, [I would prefer] social, social aspects very much.*<br>Discussion from<br>Glasgow_Workshop 3A | *“I find community for me helps me do like all sorts of things when I'm doing it with other people. And people mentioned a buddy system. But could you have some sort of little online community where people could just if they wanted to? Be accountable with other people, or encourage other people, or say what they've done, or something like that”.* |
|  |  |  | Participant_Sydney_Workshop 3 |
| **Goal setting**<br>**(Enablement)** | Customisable goals, with a minimum starting point (e.g. 1-4 bouts/day)<br>Start small and build up towards more bursts or more intense movement | *“And there has to be like basic [goal] for everybody, which is being set up and then there should be an option for you to, if you want to increase intensity or the duration or maybe just the number of times you want to do it, so there should be an option”*<br>Participant 17_Glasgow_Workshop 3B | *“I think there's gonna need to be time to set goals but then adjust the goals because some people might just find it's way too much or not enough”*<br>Participant 16_Sydney_Workshop 3 |
| **Tracking and Gamification**<br>**(Enablement and Incentivisation)** | Points systems, Real-life incentives | No explicit data in Workshop 3 | *“Maybe it could be point based? Like per minute you get a point, but then you get extra points if it's a VILPA? And you aim to get 10 points a day”*<br>Participant 16_Sydney_Workshop 3 |

**Table 6:** Intervention components suggested by participants mapped to intervention functions, the modifiable factors they address and if they were included in the final programme.

| **Intervention Function** | **Suggested Intervention Components** | **Modifiable Factor(s) Addressed** | **Included in Programme?** |
| --- | --- | --- | --- |
| Education | Information on what VILPA/MV-ILPA are | Lack of Knowledge or Clear Guidance | Yes |
|  | Information on health benefits of VILPA/MV-ILPA and movement in general | Lack of Knowledge or Clear Guidance | Yes |
| Persuasion | Emphasise brief time: benefit ratio | Perceived Energy and Effort | Yes |
|  | Emphasise that VILPA/MV-ILPA can be adapted for everyone and needs few resources | Concerns about Ability and Safety & Perceived Energy and Effort | Yes |
| Training | Guidance on what activities you could do | Concerns about Ability and Safety and Environmental Constraints | Yes |
|  | Guidance on what vigorous and moderate-vigorous intensity feels like | Concerns about Ability and Safety | Yes |
|  | Guidance on how to do VILPA/MV-ILPA safely | Concerns about Ability and Safety | Yes |
| Environmental Restructuring: Social | Sensitise the wider community to what VILPA/MV-ILPA is | Sociocultural Norms | Partly |
| Modelling | Online forums for sharing progress and encouragement | Perceived Energy and Effort | Yes |
|  | Testimonials from other community members | Concerns about Ability and Safety & Sociocultural Barriers | Yes |
|  | Buddy System or Walking Group | Perceived Energy and Effort & Sociocultural Barriers | No |
| Incentivisation | Gamification through points systems or real-life incentives | Low Mood and Mental Health | No |
| Enablement | A means to track progress either on app or using paper | Knowledge and Awareness and Low Mood and Mental Health | Yes |
|  | Use of an app or calendar to set customisable goals | Perceived Energy and Effort and Lack of Knowledge and Awareness | Yes |
|  | Recommend set goals that are accessible to everyone | Concerns about Ability and Safety & Perceived Energy and Effort | Yes |

Participants emphasised that *education and awareness* were fundamental to a feasible intervention to promote VILPA/MV-ILPA. For example, in both contexts, participants highlighted that it was important to deliver education on the concept of VILPA/MV-ILPA and the associated health benefits. However, many participants also identified the importance of education as a means to persuade others on the accessibility and adaptability of VILPA/MV-ILPA. *Social support and community building* was another key theme. Several participants in Glasgow and Sydney suggested that community-based activities can be motivating due to a sense of fun, accountability and encouragement. Participants also indicated that some way to set *goals* to do VILPA/MV-ILPA would be important-this could either be a feature of the app or a physical calendar. When asked about who should set these goals, participants in both Sydney and Glasgow suggested that there should be a small pre-set goal (e.g. 1 bout of VILPA/MV-ILPA/day) that can be adapted depending on an individual’s abilities and routine. Finally, participants in Sydney, suggested that some form *of tracking and gamification*, would also motivate them to do more VILPA/MV-ILPA. Notably, while gamification was briefly mentioned in Glasgow, this appeared to be less of a priority.

A programme theory was developed guided by previous work on the facilitators and barriers to VILPA/MV-ILPA (12–14), VILPA/MV-ILPA intervention design (15), the outcomes of the co-design workshops in Glasgow and Sydney and the behaviour change wheel (18, 19). This is depicted visually in Figure 1 and shows the relationship between modifiable factors, programme components, immediate, intermediate and long-term outcomes.

**Figure 1:**
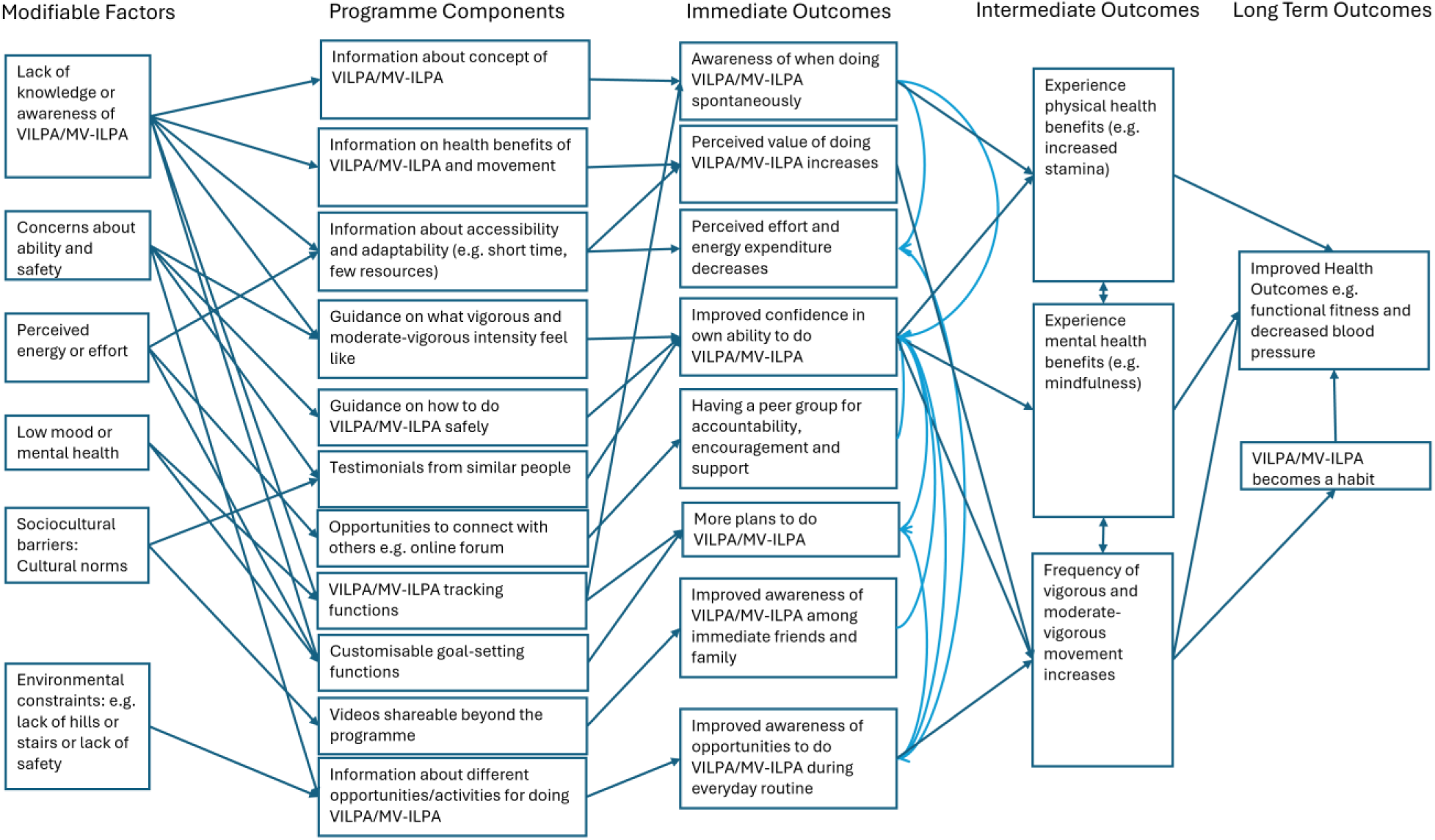
Programme theory for a co-designed intervention to promote VILPA/MV-ILPA among women from diverse communities in Sydney and Glasgow

#### Step 4. Identify how to deliver the change mechanism

From the co-design workshops were four key themes that all prioritised the accessibility of the intervention: *Delivery of Educational Materials; Tailoring and Personalisation; Digital Accessibility and App Features; and Simple and Fun Terminology.* These themes are summarised with representative quotes from Glasgow and Sydney in Table 7.

**Table 7.** Summary of themes relating to intervention delivery mechanisms including representative quotes from Glasgow and Sydney

| Theme | Examples | Representative Quote: Glasgow | Representative Quote: Sydney |
| --- | --- | --- | --- |
| **Delivery of Educational Materials** | Accessible formats including short videos, fun images or cartoons, limited text and jargon. | *“I wouldn’t want to go into reading. You know it will remind me of school. I have to read essays and words I’ll just read, then I will not complete [the intervention]”*<br>Participant 13_Glasgow_Workshop 3B | *“Videos that opens one’s eyes to the idea of short and sweet [movement] with some examples”*<br>Participant 21_Sydney_Workshop 3 |
| **Tailoring and Personalisation** | Offering choices in format (video, written info, booklet, app), social elements and language<br>Options to set and adapt personalised goals, activities and reminders | *“Yeah, that’s why we say that different formats for different audiences and then talked about apps. How they can make life easy as well social media and for the logbook for some of us who like hard copies of stuff, it can be a physical logbook or that logbook can be digital and included the app”*<br>Participant 19_Glasgow_Workshop 3B | *“So having [some information] where it’s just spoken, you know, in their language that they can understand. It is another way that can help”*<br>Participant 27_Sydney_Workshop 3 |
| **Digital Accessibility and App Features** | Free, easy-to-use apps with reminders, progress tracking, and rewards<br>Compatibility with wearables and other applications | *"If they designed the app with everything in it like all the components in one app, it would be easy for someone to use or if that app is compatible with other existing apps, that would even be better. And then we're thinking if that app is freely available"*<br>Participant 13_Glasgow_Workshop 3B | *"Needs to be free, easy to access and use, flexible and challenging enough to maintain motivation"*<br>Participant 9_Sydney_Workshop 3 |
| **Simple and Fun Terminology** | Terms that emphasise accessibility, short duration and ease of VILPA/MV-ILPA<br>Remove jargon and acronyms | *P15: That's something to write down, most of us said we liked the boosting your movement or something like that, that's why we came. It didn't have anything like VILPA or MV-ILPA in it, you tricked us [laughs]*<br>*P13: Yeah. If you had said VILPA or MV-ILPA, I wouldn't have come. I think it's not for me. Like I think it's not for me. I think it's for certain illness or athletes*<br>Discussion_Glasgow_Workshop 3B | *"I like all of the moments of movement and heart moves but terms that identify the micropattern without much explanation such as quick fitness moves, bite sized, mini moves / bursts is easier for newbies to grasp"*<br>Participant 11_Sydney_Workshop 3 |

The accessible *delivery of the educational materials* was a priority across groups in Glasgow and Sydney. Participants emphasised the need for fun, simple and engaging information, preferably in visual formats (e.g. short videos and pictures/cartoons) as written text could be viewed as unappealing. In both contexts, participants also suggested that educational materials should cite credible sources such as the University of Sydney and University of Glasgow, to highlight the legitimacy of the study amidst a wealth of physical activity misinformation online.

Finally, although videos with optional transcripts appeared to be the preferred format for most participants, physical logbooks or information booklets would also be welcomed for those who are less comfortable with technology.

Indeed, a common thread across delivery mechanisms was the need for *tailoring and personalisation.* This encompassed the inclusion of different formats of information, different pathways through the app, a culturally inclusive list of activities and personalisable goals and reminders. The importance of adaptations for different age groups, cultures, languages, physical abilities and learning styles were also highlighted in Sydney and Glasgow. Although discussions were kept broad to allow for a range of delivery mechanisms; there was a focussed emphasis on *digital accessibility*, specifically the delivery of the intervention through a mobile-phone app. When considering this approach, participants emphasised that the app would need to be free at the point of use, simple and easy to use, be compatible with other apps and wearable devices and include multiple different features.

Finally, participants in Glasgow and Sydney were invited to reflect on and provide suggestions for new terminology over the course of the Workshops. Participants emphasised that the use of acronyms should be kept to a minimum, and jargon, including the word ‘*vigorous*’, should be replaced, with simple, appealing terms. In Sydney and Glasgow participants suggested terms such as “*mindful”* or “*manageable movement”* to centre the accessibility of VILPA/MV-ILPA. The short time duration of VILPA and MV-ILPA was also important. For example, in Sydney participants suggested the term “*mini moves*” and in Glasgow participants suggested “*short bursts*” or “*snippets*”. Several participants in Glasgow Workshop 3 highlighted that the language used in recruitment: *Boosting Everyday Movement*, had already been successful at getting people involved in the study. Participant 15 went a step further to suggest that the term could be adapted to apply directly to the acronyms VILPA and MV-ILPA.

> “So, you could be boosted and super boosted, because you’ve got boosting your movement. So, you could be boosted, so you’re doing more breathing and then super boosted that you’re out for a… I don’t know…But this is what’s attracted us”

Participant 15_Glasgow_Workshop 3B

Participant 15 suggested that the moderate-vigorous version MV-ILPA could be known simply as a “*Boost”* and that VILPA could be known as a “*Super Boost”*, with the “*super”* implying even more effort is required to achieve it. These became the working terms for describing VILPA and MV-ILPA in the intervention.

### Final Intervention

Upon analysis of the findings from co-design process in Glasgow and Sydney a 6-week intervention programme was developed that involved educational materials, VILPA/MV-ILPA tracking and goal setting. Due to the popularity of social components and mixed perceptions of the accessibility of digital technology, the intervention was designed with three different delivery mechanisms which could be tested in the next phase. These included: 1) the use of the smartphone application only; 2) the smartphone application and a wearable device; and 3) two in-person workshops, the use of a mobile phone app and a wearable device; By exploring these three delivery mechanisms, the feasibility of different tools to track VILPA/MV-ILPA as well as how much additional social contact and support is necessary to promote sustainable behaviour change could be explored. The final intervention components and delivery mechanisms for each week and group are summarised in Table 8. On the smartphone application, educational modules were designed to be released weekly, and participants could refer back to previous weeks at any time, other functions including the online forum, tracking (linked to phone step data or wearable device) and goal setting were available at any time from Week 1. Additionally, in response to participant perspectives on accessibility, the educational materials were presented using bright, fun colours, images and cartoons, simple language and short videos and text.

**Table 8:**
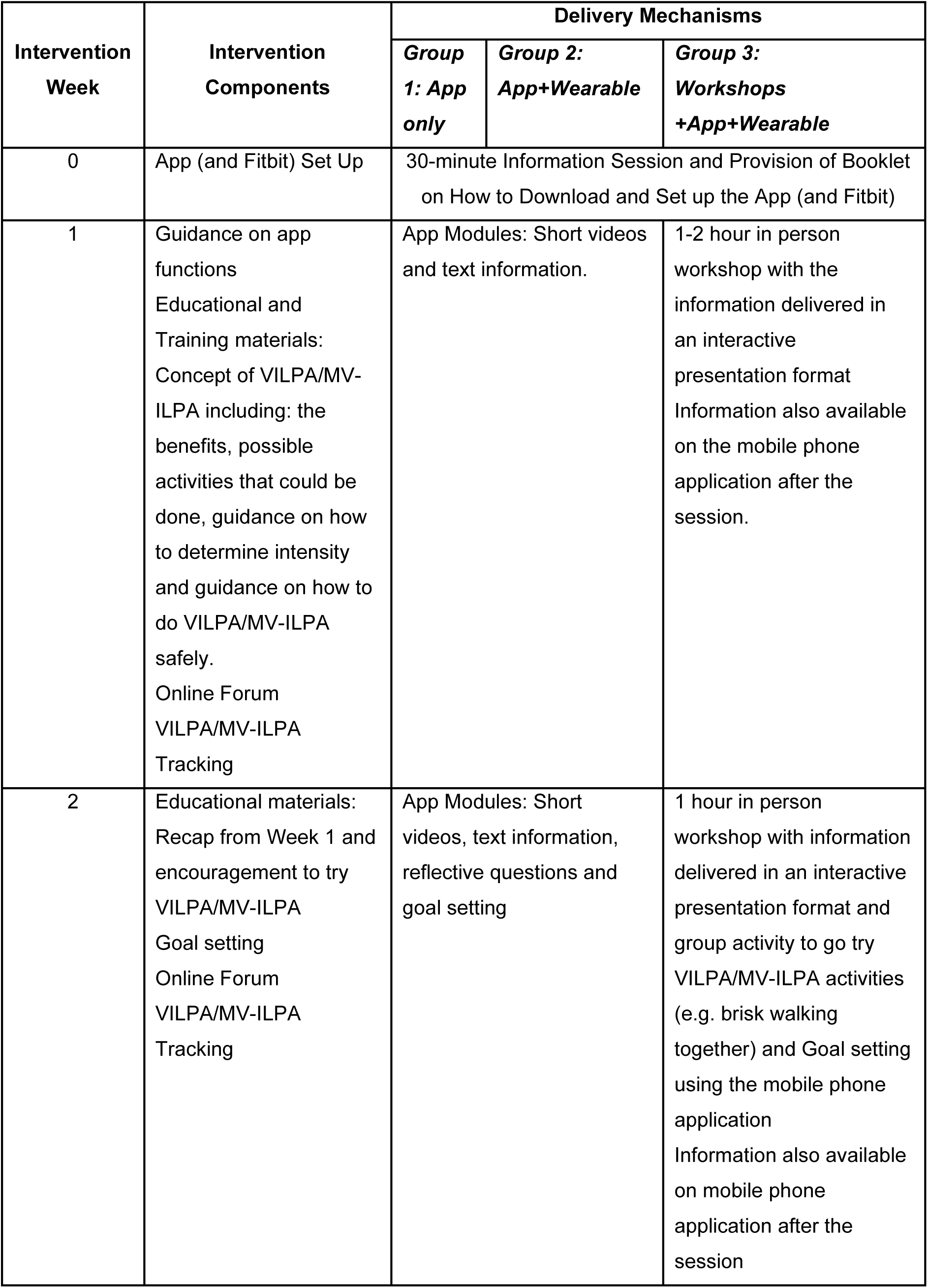
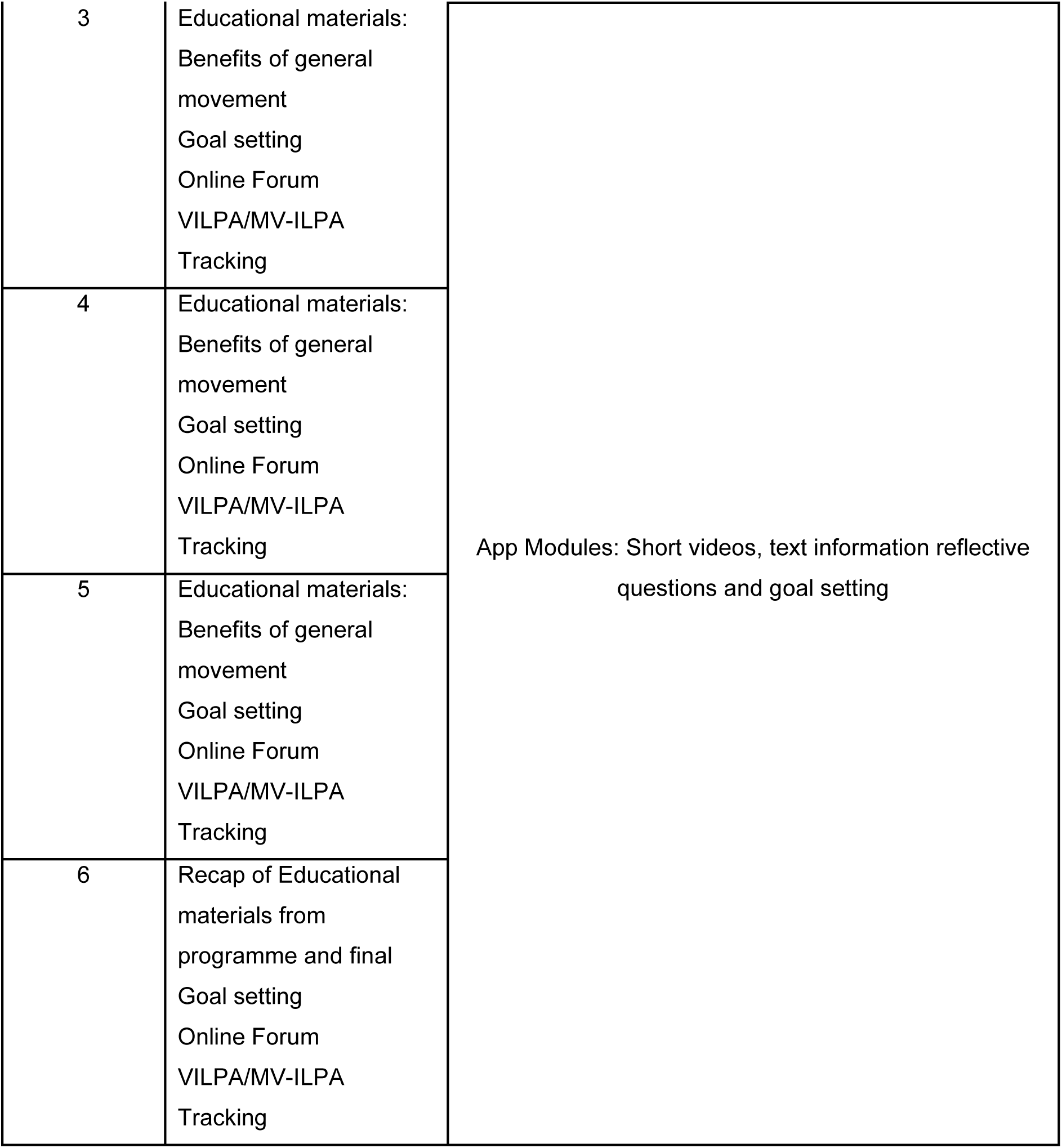
Final 6-week intervention components and delivery mechanisms

| Intervention Week | Intervention Components | Delivery Mechanisms |  |  |
| --- | --- | --- | --- | --- |
|  |  | Group 1: App only | Group 2: App+Wearable | Group 3: Workshops +App+Wearable |
| 0 | App (and Fitbit) Set Up | 30-minute Information Session and Provision of Booklet on How to Download and Set up the App (and Fitbit) |  |  |
| 1 | Guidance on app functions<br>Educational and Training materials: Concept of VILPA/MV-ILPA including: the benefits, possible activities that could be done, guidance on how to determine intensity and guidance on how to do VILPA/MV-ILPA safely.<br>Online Forum<br>VILPA/MV-ILPA Tracking | App Modules: Short videos and text information. |  | 1-2 hour in person workshop with the information delivered in an interactive presentation format<br>Information also available on the mobile phone application after the session. |
| 2 | Educational materials: Recap from Week 1 and encouragement to try VILPA/MV-ILPA<br>Goal setting<br>Online Forum<br>VILPA/MV-ILPA Tracking | App Modules: Short videos, text information, reflective questions and goal setting |  | 1 hour in person workshop with information delivered in an interactive presentation format and group activity to go try VILPA/MV-ILPA activities (e.g. brisk walking together) and Goal setting using the mobile phone application<br>Information also available on mobile phone application after the session |

|  |  |  |
| --- | --- | --- |
| 3 | Educational materials:<br>Benefits of general movement<br>Goal setting<br>Online Forum<br>VILPA/MV-ILPA<br>Tracking | App Modules: Short videos, text information reflective questions and goal setting |
| 4 | Educational materials:<br>Benefits of general movement<br>Goal setting<br>Online Forum<br>VILPA/MV-ILPA<br>Tracking |  |
| 5 | Educational materials:<br>Benefits of general movement<br>Goal setting<br>Online Forum<br>VILPA/MV-ILPA<br>Tracking |  |
| 6 | Recap of Educational materials from programme and final<br>Goal setting<br>Online Forum<br>VILPA/MV-ILPA<br>Tracking |  |

## Discussion

This study presents the co-design of a theory-informed intervention to promote Vigorous Intermittent Lifestyle Physical Activity (VILPA) and Moderate-Vigorous Intermittent Lifestyle Physical Activity (MV-ILPA) among socioeconomically diverse women in Glasgow and Sydney. Through participatory co-design workshops, participants shaped the development of an intervention involving key educational components, training, goal setting and tracking to promote VILPA/MV-ILPA. These components lend themselves to the use of digital interventions such as mobile phone applications and wearable devices which could be used promote VILPA/MV-ILPA (20, 33). However, participants also emphasised the importance of social elements as means to provide encouragement and accountability. Social interventions have been shown to be less cost effective than less intensive forms e.g. digital interventions (34). Therefore, social components such as workshops could be used in combination with digital technology to deliver the intervention by providing both social support and tracking (20) and be a more cost-effective solution than providing in-person workshops alone (34). This is something that will need to be tested further in the next phase of intervention development.

The findings also offer several key insights into the barriers and facilitators to integrating VILPA/MV-ILPA into everyday life for women in Glasgow and Sydney. Participants highlighted key barriers including ageing and physical limitations, concerns over their ability to do vigorous activity safely, environmental and sociocultural constraints. The facilitators, including the adaptability of VILPA and positive perspectives on the idea of dual-purpose activities suggest that the accessibility of the concept is highly promising to promote more movement among groups who do little physical activity. Notably, the facilitators and barriers to VILPA/MV-ILPA identified in the present study largely mirrored previous work in Australia suggesting that, with appropriate care and attention to specific sociocultural barriers, interventions to promote VILPA/MV-ILPA could be readily adapted to different contexts and groups (12, 14, 15).

This study also emphasised the importance of accessibility in the design and delivery of interventions. This includes how information is presented, keeping the intervention fun and enjoyable, and ensuring it can be adapted to different groups and their unique needs. While some of these points are easy to address in the initial development, adaptions such as multiple languages and display formats should be considered in future iterations. Finally, the co-created terminology such as “Boost” and “Super Boost” could be highly promising to make the concept of VILPA/MV-ILPA more accessible and promote engagement in an intervention.

A key innovation of this study lies in its use of co-design to develop an intervention that is both contextually grounded and theoretically robust. Through workshops that promoted active collaboration with women of diverse socioeconomic backgrounds to not only gain their perspectives on VILPA/MV-ILPA but also draw on the Behaviour Change Wheel (18, 19), this intervention is both tailored to real-world experiences and critically informed by behaviour change theory. The development of the intervention is also guided by the 6SQuID model, and the MRC framework for complex interventions (16, 17, 25) and the development of a programme theory support the feasibility of the intervention.

### Strengths and Limitations

The study had several strengths and limitations. Strengths included its international scope, theory-informed design, and deep engagement with community members from diverse socioeconomic backgrounds. The use of participatory methods across multiple sites allowed for the generation of broad and context-specific insights. There were also several limitations, for example, the delivery of the workshops needed to be adapted to the specific context, meaning that two sets of workshops were run in Glasgow with smaller groups and one larger hybrid workshop was run in Sydney, where most participants were online. These differences may have contributed to different interactions between participants and facilitators, differences in the breadth of discussion and slight changes in the protocols. Furthermore, the programme theory was not formally validated with participants or other stakeholders.

## Conclusion

This study demonstrates the value of co-design in developing a contextually grounded and theory-informed intervention to promote VILPA and MV-ILPA among socioeconomically diverse women. The findings highlight the importance of tailoring VILPA/MV-ILPA interventions to the lived experiences of target populations. Future research should focus on feasibility and pilot testing across the three proposed delivery mechanisms, with particular attention to digital accessibility and social support. Iterative refinement of the programme theory and terminology will be essential to ensure sustained engagement.

## Data Availability

All qualitative data relating to the findings are included in the manuscript in the form of quotations.

## Acknowledgements

We thank the extensive involvement of community members from Australia and the UK involved in the co-design of this intervention.

